# An update to the LIFEtime-perspective CardioVascular Disease (LIFE-CVD) model for prediction of individualized lifetime benefit from cardiovascular risk factor management in apparently healthy people

**DOI:** 10.1101/2021.03.15.21253400

**Authors:** Tamar I. de Vries, Nicole E.M. Jaspers, Frank L.J. Visseren, Jannick A.N. Dorresteijn

## Abstract

**Introduction:** The previously developed LIFEtime-perspective CardioVascular Disease (LIFE-CVD) model can be used to predict lifetime cardiovascular disease risk, CVD-free life expectancy, and lifetime benefit from cardiovascular risk factor treatment in apparently healthy people aged 45 to 80 years. However, there was an unmet need to be able to apply the model in patients younger than 45 years, and to accurately estimate treatment effects in patients with a life expectancy exceeding 90 years.

**Aim:** Update the LIFE-CVD model to enable application of the model in people aged 35 to 89 years, and to allow more accurate estimation of treatment effects in patients with a life expectancy exceeding 90 years.

**Methods:** The study was conducted using data from the same studies as were used for derivation and validation of the original model, including the Multi-Ethnic Study of Atherosclerosis (MESA) cohort, Atherosclerosis Risk in Communities Study (ARIC) cohort, and the European Prospective Investigation into Cancer-Netherlands (EPIC-NL) and EPIC-Norfolk cohort studies. Age-specific baseline survivals were smoothed by predicting the progression of baseline survivals with age, using a local polynomial regression function and a exponential function for CVD, and non-CVD mortality baseline survivals respectively. Using these functions, baseline survivals were then extrapolated to the age range of 35 to 100 years. External validation using the newly updated baseline survivals was performed.

**Results:** Performance of the updated model was not dissimilar from the original model, with C-statistics for discrimination ranging from 0.70-0.76 in the external study populations. Calibration plots showed a good agreement between predicted and observed 10-year CVD risks. Estimation of treatment effects in patients with a life expectancy exceeding 90 years was improved.

**Conclusion:** This update of the LIFE-CVD model improves the clinical usability of the model by increasing the age range and improving the method of estimation of lifetime treatment effects.

## Introduction

In 2019, the LIFEtime-perspective CardioVascular Disease (LIFE-CVD) model for the prediction of lifetime cardiovascular disease risk, life expectancy free from CVD, and lifetime treatment benefit from cardiovascular risk factor treatment in apparently healthy people aged 45 to 80 years was published in the European Heart Journal.^1^ This risk model is available as online tool on the website www.U-Prevent.com. Using the model in daily clinical practice, treatment effects estimated in patients with a life expectancy exceeding 90 years appeared to be underestimated. Furthermore, there was an unmet need to be able to apply the model in patients younger than 45 years. The present paper aims to solve both these issues by providing an updated list of age-specific baseline survival rates for a wider age-range and describe an adapted method for lifetime benefit calculation.

## Methods

### Study populations

The study was conducted using data from the same studies as described in the original paper by Jaspers et al.,^1^ including the Multi-Ethnic Study of Atherosclerosis (MESA) cohort,^2^ Atherosclerosis Risk in Communities Study (ARIC) cohort,^3^ and the European Prospective Investigation into Cancer-Netherlands (EPIC-NL) and EPIC-Norfolk cohort studies.^4,5^ Participants between 35 and 45 years of age or aged >80 years, which had been excluded from original model derivation, were included in the present update.

### The LIFE-CVD prediction algorithm

Details on the methods for derivation and validation of the LIFE-CVD algorithm have been described previously.^1^ In summary, the LIFE-CVD prediction algorithm exists of two complementary externally validated competing-risk adjusted left-truncated Fine and Gray-hazard functions for lifetime predictions for major cardiovascular events (MACE; myocardial infarction, stroke, or cardiovascular mortality) and non-cardiovascular death in apparently healthy persons.^1^ It was derived in the MESA cohort (n=6715) and validated in the ARIC, HNR, EPIC-NL and EPIC-Norfolk studies (total n=62808), with C-statistics for discrimination ranging between 0.67-0.76. It is based on the following clinically readily available characteristics: sex, systolic blood pressure, non-high density lipoprotein cholesterol, body mass index, smoking status, diabetes mellitus, and parental history of premature myocardial infarction. The model can be used for estimation of both 10-year risk of MACE and CVD-free life expectancy. Additionally, it can be used to predict the lifetime treatment effects from cholesterol lowering, blood pressure lowering, antithrombotic therapy, and smoking cessation.

Individual estimations are based on lifetables with 1-year age intervals. Each life-year has an age-specific 1-year baseline survival for both CVD-events and for the competing outcome of non-CVD mortality. By combining the baseline survival with the clinical predictors (coefficients), the individual risk of having a CVD-event or dying from non-cardiovascular causes can be estimated for each life-year. The cumulative survival for each life-year can then be estimated by multiplying the survival probability of that life year (1 minus CVD-risk minus non-CVD mortality risk) with the survival probability at the beginning of each life-year. This process is repeated until the maximum model age. The CVD-free life expectancy is defined as the median survival without a CVD-event or death (i.e. the age at which the cumulative survival probability becomes <50%).

### Updating the LIFE-CVD algorithm

The age-specific baseline survivals for the original LIFE-CVD prediction algorithm (presented in Supplemental Table 3 of the 2019 paper, and Table 1 of this report)^1^ are based on the observed events per life-year (i.e. at which age the observed events occurred). Due to chance, there is some variation between life years that cannot be explained by the natural progression of the 1-year risk for CVD or non-CVD mortality with increasing age. By predicting the progression of baseline survivals with age, the corresponding individual survival plots will also be smoothed and therefore more intuitive than when using the observed baseline survivals. Additionally, by using a model to predict the progression of baseline survivals with age, rather than using the observed age-specific baseline survivals, it becomes possible to extrapolate the baseline survivals for ages outside of this original age range according to the formula predicting the baseline survivals.

**Table 1:** Original age-specific baseline survival for CVD-events and non-CVD mortality based on the observed events per life-year

|  | 1-year CVD<br>baseline<br>survival | 1-year non-CVD<br>mortality baseline<br>survival |  | 1-year CVD<br>baseline<br>survival | 1-year non-CVD<br>mortality baseline<br>survival |
| --- | --- | --- | --- | --- | --- |
| Age |  |  | Age |  |  |
| 45 | 1 | 1 | 68 | 0.999645771 | 0.970398098 |
| 46 | 1 | 0.979380684 | 69 | 0.999663492 | 0.965410198 |
| 47 | 0.999725398 | 0.985480904 | 70 | 0.999566041 | 0.956805621 |
| 48 | 1 | 0.978988162 | 71 | 0.999513256 | 0.954653450 |
| 49 | 1 | 1 | 72 | 0.999692656 | 0.955233741 |
| 50 | 0.999878410 | 0.98607217 | 73 | 0.999702516 | 0.953443559 |
| 51 | 0.999516952 | 0.988522333 | 74 | 0.999679667 | 0.969421315 |
| 52 | 0.999591619 | 0.985415629 | 75 | 0.999647779 | 0.947674229 |
| 53 | 0.999792279 | 0.979056609 | 76 | 0.999630632 | 0.939724478 |
| 54 | 0.999879900 | 0.985072499 | 77 | 0.999686488 | 0.954401682 |
| 55 | 0.999412090 | 0.989896930 | 78 | 0.999661579 | 0.930794801 |
| 56 | 0.999571482 | 0.990764307 | 79 | 0.999598915 | 0.947523700 |
| 57 | 0.999699809 | 0.985900751 | 80 | 0.999725086 | 0.944907534 |
| 58 | 0.999682114 | 0.986623736 | 81 | 0.999769337 | 0.931903316 |
| 59 | 0.999511434 | 0.987138590 | 82 | 0.999684311 | 0.918651265 |
| 60 | 0.999322636 | 0.979874870 | 83 | 0.999603926 | 0.916640537 |
| 61 | 0.999656409 | 0.973393148 | 84 | 0.999668146 | 0.882636308 |
| 62 | 0.999481427 | 0.966878497 | 85 | 0.999583733 | 0.902748240 |
| 63 | 0.999630190 | 0.985990456 | 86 | 0.999488751 | 0.896307118 |
| 64 | 0.999465298 | 0.977289395 | 87 | 0.999585936 | 0.884407038 |
| 65 | 0.999726810 | 0.964981886 | 88 | 0.999723251 | 0.918574710 |
| 66 | 0.999772552 | 0.976599292 | 89 | 0.999513316 | 0.868249158 |
| 67 | 0.999693501 | 0.967697361 |  |  |  |

The updated baseline survivals were predicted according to functions weighted for the number of individual participants contributing data to each life-year. The CVD baseline survivals were predicted and smoothed using local polynomial regression (function *loess*, package stats in R studio) using a smoothing parameter α of 1.05. The non-CVD mortality baseline survivals followed an exponential function according to the form *E(Y) = a * exp(bx) + c* and were predicted using a non-linear regression function (Figure 1). The baseline survivals were then extrapolated to the age range of 35 to 100 years, allowing predictions over a wider age range (Table 2).

**Table 2:** Updated age-specific baseline survival for CVD-events and non-CVD mortality

|  | 1-year CVD<br>baseline<br>survival | 1-year non-CVD<br>mortality baseline<br>survival |  | 1-year CVD<br>baseline<br>survival | 1-year non-CVD<br>mortality baseline<br>survival |
| --- | --- | --- | --- | --- | --- |
| Age |  |  | Age |  |  |
| 35 | 0.99991823 | 0.99307706 | 68 | 0.99963267 | 0.96820266 |
| 36 | 0.99990882 | 0.99289348 | 69 | 0.99963845 | 0.96615015 |
| 37 | 0.99989875 | 0.99269573 | 70 | 0.99964348 | 0.96394804 |
| 38 | 0.99988803 | 0.99248272 | 71 | 0.99964759 | 0.96158622 |
| 39 | 0.99987669 | 0.99225328 | 72 | 0.99965088 | 0.95905400 |
| 40 | 0.99986477 | 0.99200614 | 73 | 0.99965348 | 0.95634012 |
| 41 | 0.99985230 | 0.99173997 | 74 | 0.99965548 | 0.95343274 |
| 42 | 0.99983936 | 0.99145329 | 75 | 0.99965695 | 0.95031941 |
| 43 | 0.99982601 | 0.99114455 | 76 | 0.99965794 | 0.94698712 |
| 44 | 0.99981233 | 0.99081206 | 77 | 0.99965847 | 0.94342224 |
| 45 | 0.99979842 | 0.99045401 | 78 | 0.99965857 | 0.93961058 |
| 46 | 0.99978436 | 0.99006846 | 79 | 0.99965824 | 0.93553740 |
| 47 | 0.99977027 | 0.98965332 | 80 | 0.99965748 | 0.93118742 |
| 48 | 0.99975627 | 0.98920634 | 81 | 0.99965629 | 0.92654486 |
| 49 | 0.99974247 | 0.98872513 | 82 | 0.99965466 | 0.92159349 |
| 50 | 0.99972899 | 0.98820709 | 83 | 0.99965258 | 0.91631671 |
| 51 | 0.99971594 | 0.98764944 | 84 | 0.99965001 | 0.91069758 |
| 52 | 0.99970343 | 0.98704922 | 85 | 0.99964692 | 0.90471895 |
| 53 | 0.99969156 | 0.98640322 | 86 | 0.99964327 | 0.89836352 |
| 54 | 0.99968043 | 0.98570803 | 87 | 0.99963904 | 0.89161400 |
| 55 | 0.99967011 | 0.98495998 | 88 | 0.99963418 | 0.88445322 |
| 56 | 0.99966066 | 0.98415513 | 89 | 0.99962867 | 0.87686428 |
| 57 | 0.99965211 | 0.98328928 | 90 | 0.99962247 | 0.86883075 |
| 58 | 0.99964450 | 0.98235793 | 91 | 0.99961554 | 0.86033685 |
| 59 | 0.99963787 | 0.98135627 | 92 | 0.99960784 | 0.85136764 |
| 60 | 0.99963223 | 0.98027914 | 93 | 0.99959933 | 0.84190928 |
| 61 | 0.99962760 | 0.97912105 | 94 | 0.99958995 | 0.83194926 |
| 62 | 0.99962404 | 0.97787613 | 95 | 0.99957966 | 0.82147664 |
| 63 | 0.99962160 | 0.97653813 | 96 | 0.99956838 | 0.81048235 |
| 64 | 0.99962042 | 0.97510036 | 97 | 0.99955607 | 0.79895946 |
| 65 | 0.99962066 | 0.97355572 | 98 | 0.99954263 | 0.78690343 |
| 66 | 0.99962260 | 0.97189666 | 99 | 0.99952800 | 0.77431242 |
| 67 | 0.99962652 | 0.97011515 |  |  |  |

**Figure 1:**
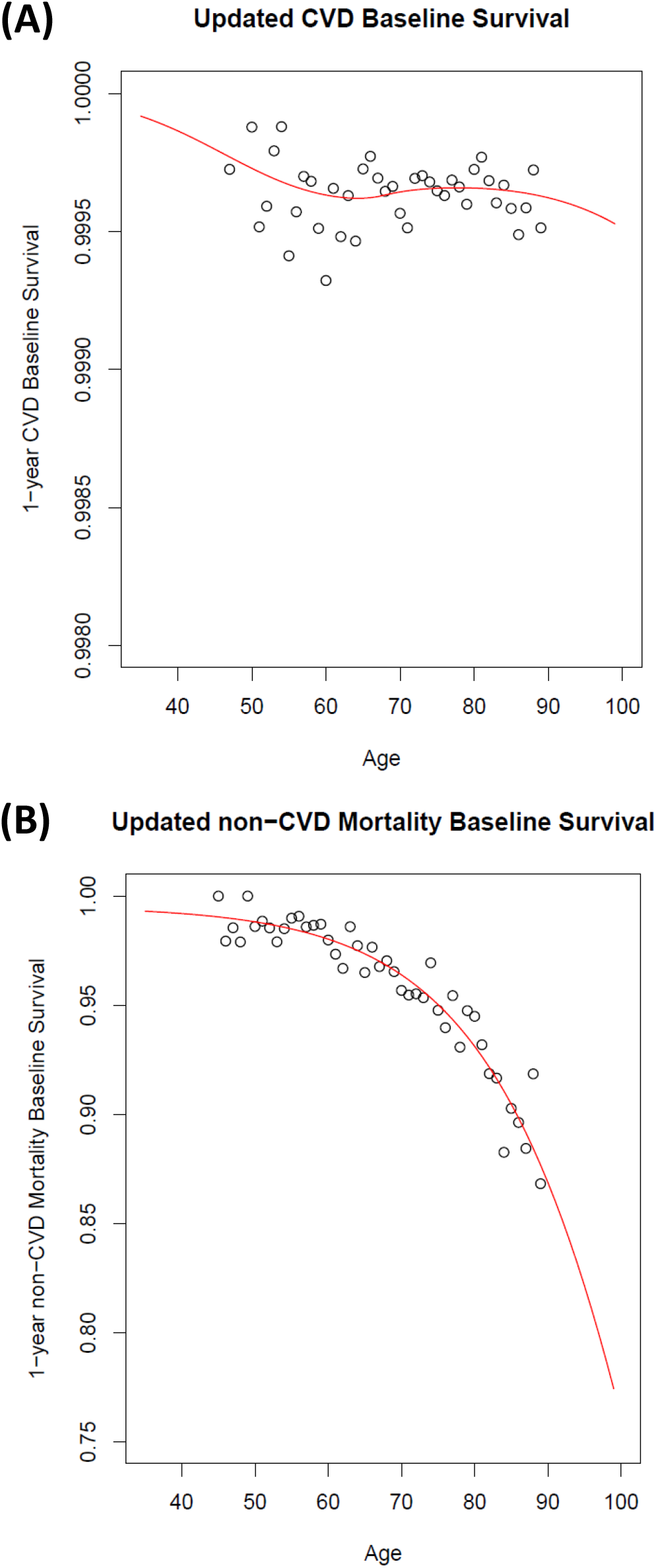
Smoothing and extrapolation of baseline survivals for (A) 1-year CVD baseline survival (using local polynomial regression), and (B) 1-year non-CVD mortality baseline survival (using non-linear regression). Black dots indicate the original baseline survivals based on the observed events per life-year, the red lines show the predicted progression of baseline survivals from the age of 35 years to 100 years (the updated baseline survivals).

Finally, external validation using the newly updated baseline survivals was then performed in ARIC, HNR, EPIC-NL, and EPIC-Norfolk. Model discrimination was assessed using c-statistics and agreement between expected-and-observed 10-year risk was assessed visually using calibration plots.

### Calculating treatment effects when the life expectancy exceeds 100 years

Treatment benefit for each risk factor treatment is estimated as the difference between on- and off-treatment median CVD-free life expectancy. In people whose life expectancy exceeds the model’s maximum age, this approach cannot be used as the cumulative survival curve does not drop below 50%. Previously, we proposed using the difference in area under the curve (AUC) in such cases as a possible solution. However, the AUC-method gives underestimation of true lifetime treatment benefit. As a better alternative, we here propose a new method of using the last observed cumulative survival. This means that in the case the on-treatment CVD-free cumulative survival exceeds 50% at the maximum age (i.e. 100 years), lifetime treatment benefit is defined as the difference between the maximum age and the age with the corresponding predicted percentage off-treatment cumulative survival.

## Results

The updated age-specific baseline survivals are presented in Table 2. Internal and external validation of the model using the updated baseline survival and the wider age range are shown in Figure 2. The model performance of the updated model was not dissimilar from model performance as presented in the 2019 paper (Figure 2). C-statistics for discrimination in the external study populations range from 0.70-0.76.

**Figure 2:**
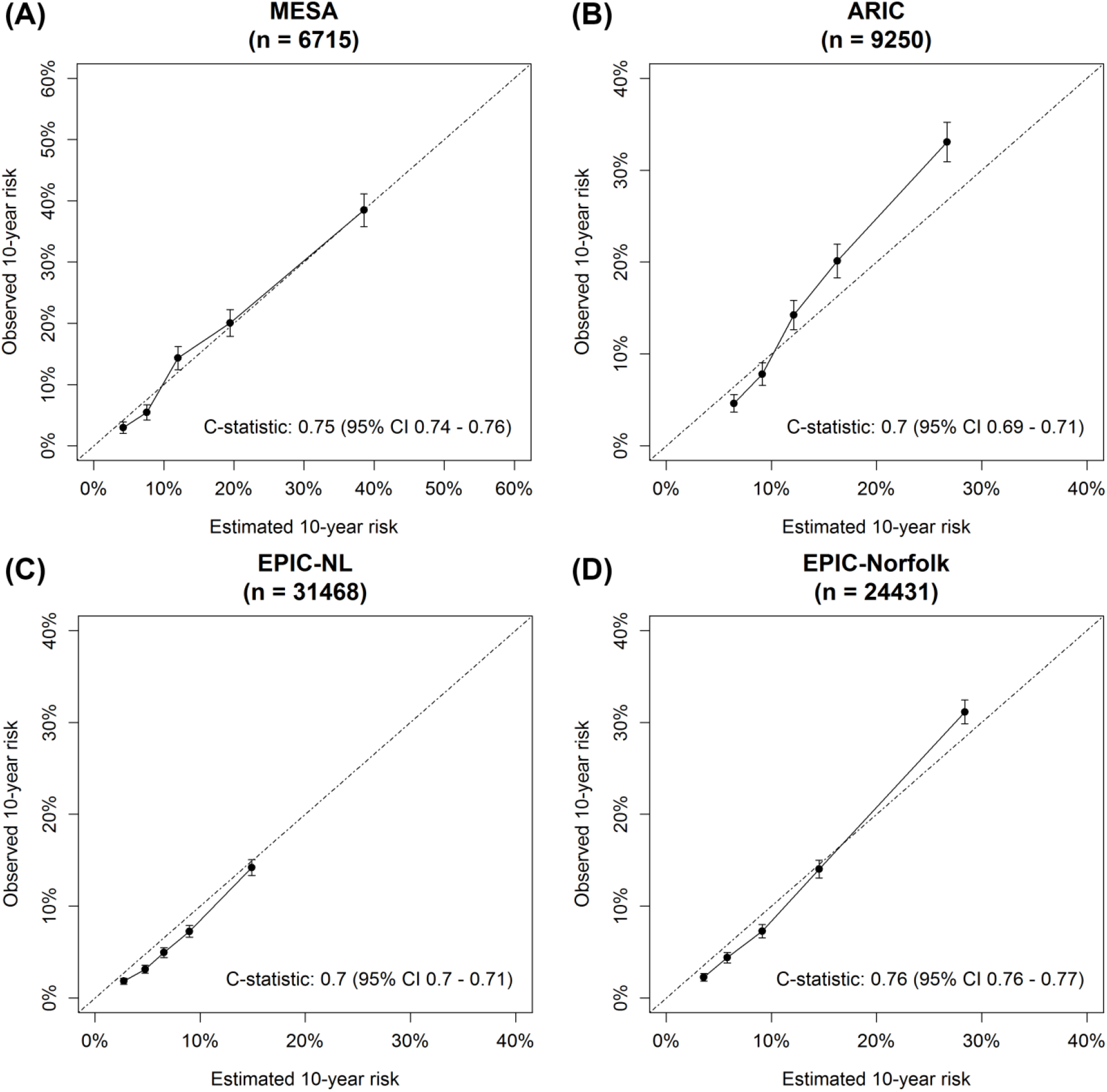
Calibration plots of expected versus observed risks with C-statistics for discrimination of the updated model stratified for age groups: (A) internal validation in MESA, (B) external validation in ARIC, (C) external validation in EPIC-NL, and (D) external validation in EPIC-Norfolk

To show an example of how the updated methodology influences the treatment benefit estimations, Figure 3 shows the distribution of treatment effects (using an example of statin treatment) comparing the original methodology to the updated methodology. The updated methodology gives noticeably higher mean treatment effects, as the method based on the AUC gave important underestimation of treatment effect in all patients with a median life expectancy higher than 90 years.

**Figure 3:**
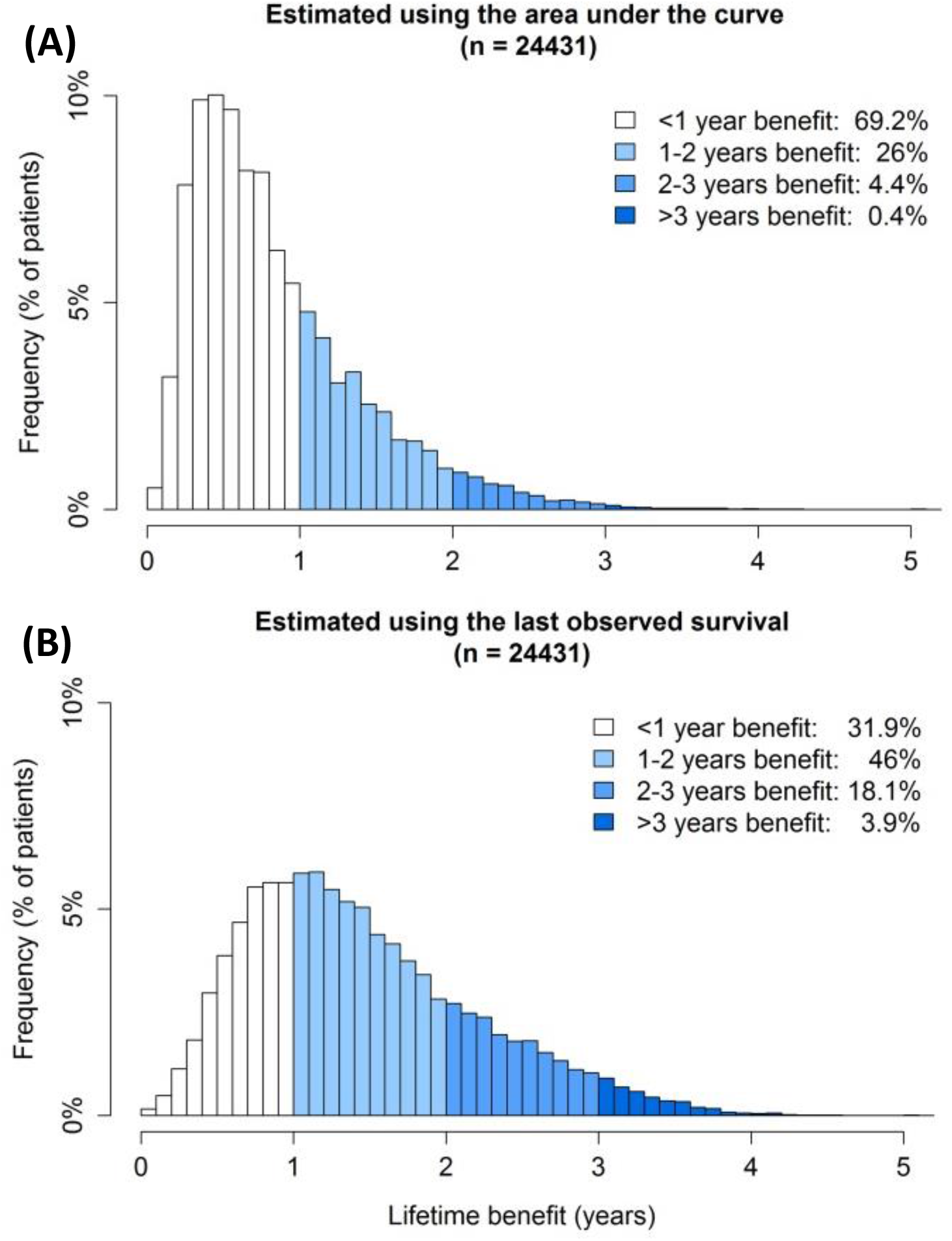
Distribution of treatment effects in the EPIC-Norfolk study population of a hypothetical situation where moderate-intensity lipid lowering therapy (e.g. simvastatin 40 mg) is initiated in all participants, using the area under the curve when life expectancy exceeds the model’s maximum age (A) compared to using the last observed survival (B)

Figure 4 shows an individual person example comparing the original methodology to the updated methodology, by showing an individual survival curve for a currently 65-year old patient. Figure 4b, the updated model, shows that the new model gives a smoother curve that extends to 100 years. Because in Figure 4a the survival curve did not drop below 50% and the treatment effect was estimated using the difference in AUC, the treatment effect was estimated as 0.3 years. In Figure 4b, however, the survival curve did drop below 50% and the lifetime treatment effect was estimated as 1.1 years (the difference between 91.6 and 92.7 years). In a third scenario, where the age range of the model was not extended to 90, but the last know survival was used for estimating the treatment effect, the treatment effect for this patient would have been 1.0 years (not shown in the figure), much closer to the 1.1 years estimated when the age range was extended. Both figures 3 and 4 thus demonstrate how the AUC underestimates treatment effect compared to using the last observed survival.

**Figure 4:**
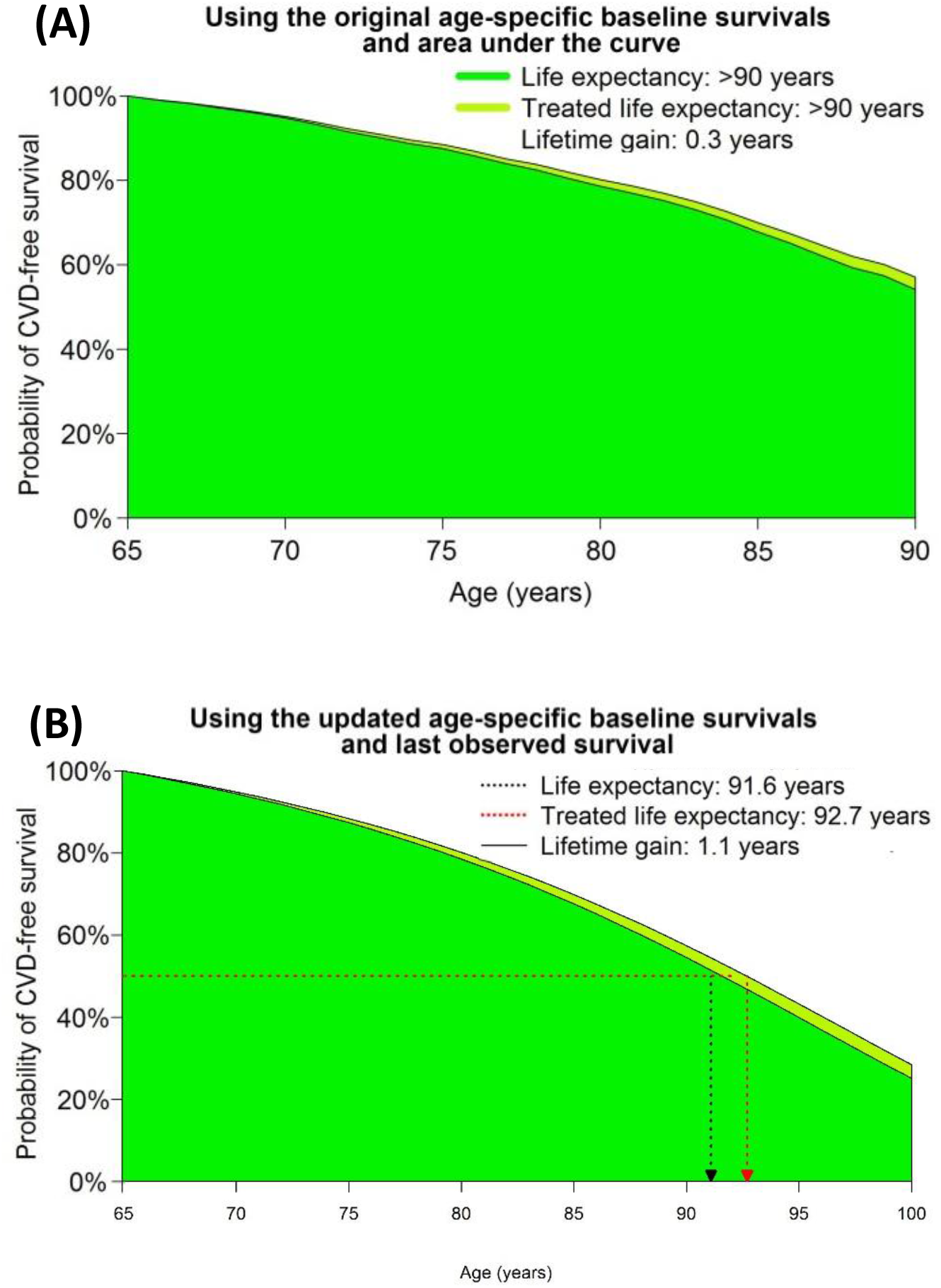
an individual person example of a survival curve for predicting the treatment effect from moderate-intensity lipid lowering therapy, using the original methodology based on difference in area under the curve (A) to the updated methodology using the last observed survival (B).

## Discussion

In this update of the LIFE-CVD model for the prediction of lifetime cardiovascular disease risk, life expectancy free from CVD, and lifetime treatment benefit from cardiovascular risk factor treatment in apparently healthy people, we have shown that it is possible to extend the age range of this lifetime prediction model without comprising on model performance. Furthermore, we have developed a new method to predict treatment effect of cardiovascular risk factor treatment in persons with a high median life expectancy.

Traditionally, in international guidelines 10-year risk models are used to decide which patients to treat with (pharmacological) cardiovascular risk factor lowering therapies.^6,7^ However, during the last decades, 10-year risk of especially fatal CVD has decreased importantly in all age groups in the Western world.^8^ Keeping the risk thresholds for treatment the same, less and less people would be treated with preventive medication. However, especially in the older age range, CVD becomes more and more prevalent.

As atherosclerosis is a lifelong gradual and progressive process, it seems logical to intervene early in that process to slow it down or stop it from happening all together. Younger patients with a high burden of cardiovascular risk factors may have a high lifetime risk but a very low 10-year risk. Based on 10-year risk thresholds for initiation of risk factor treatment in the current international guidelines,^6,7^ younger apparently healthy persons are usually not eligible for preventive pharmacotherapy. The updated LIFE-CVD model, which now allows lifetime risk and lifetime treatment benefit estimation starting at the age of 35 years, can help in selecting patients who will benefit from risk factor treatment at a young age. Perhaps even more importantly, lifetime risk and treatment effect estimations can facilitate in communicating the importance of healthy lifestyle and risk factor management at a young age, especially smoking cessation, even when pharmacotherapy is not (yet) considered.

The updated LIFE-CVD model has as advantage that is gives more accurate lifetime estimations of treatment effects in *all* patients, even those who have life expectancies exceeding 90 years. As the worldwide life expectancy keeps increasing,^9^ this will be increasingly important.

Some limitations of this study need to be acknowledged. Firstly, as this is an update of the original LIFE-CVD model, this model suffers from the same limitations as the original model. The discussion of these limitations can be found in the original 2019 paper by Jaspers et al.^1^ Secondly, the used methodology assumes that the age-specific baseline survival follows a certain line that can be predicted, and more importantly, can be extrapolated outside of the original age range. Looking at the plot of the original baseline survival (in the supplemental material) we do not have a reason to assume that this assumption is not met. Furthermore, the extrapolation does not seem to influence validation. However, we cannot know for certain whether this is actually true, as there are too little persons aged < 45 or >90 in the study population to formally assess this assumption.

In conclusion, this update to the LIFE-CVD algorithms improves the clinical usability of the model by increasing the age range and improving the method of estimation of lifetime treatment effects.

## Data Availability

Data requests may be send to the corresponding author.

